# Monocyte to Lymphocyte ratio is highly specific in diagnosing latent tuberculosis and declines significantly following tuberculosis preventive therapy: *a cross-sectional and nested prospective observational study*

**DOI:** 10.1101/2023.05.02.23289422

**Authors:** Jonathan Mayito, David B. Meya, Akia Miriam, Flavia Dhikusooka, Joshua Rhein, Christine Sekaggya-Wiltshire

## Abstract

Interferon-gamma release assay and tuberculin skin test use is limited by costly sundries and cross-reactivity with non-tuberculous mycobacteria and Bacille Calmette-Guérin (BCG) vaccination respectively. We investigated the Monocyte to Lymphocyte ratio (MLR) as a biomarker to overcome these limitations and for use in monitoring response to tuberculosis preventive therapy (TPT). We conducted a cross-sectional and nested prospective observational study among asymptomatic adults living with Human Immuno-deficiency Virus (HIV) in Kampala, Uganda. Complete blood count (CBC) and QuantiFERON-TB® Gold-plus were measured at baseline and CBC repeated at three months. Multivariate logistic regression was performed to identify factors associated with a high MLR and decline in MLR. We recruited 110 adults living with HIV and on antiretroviral therapy, of which 82.5% (85/110) had suppressed viral loads, 71.8% (79/110) were female, and 73.6% (81/110) had a BCG scar. The derived MLR diagnostic cut-off was 0.35, based on which the MLR sensitivity, specificity, positive predictive value, and negative predictive value were 12.8%, 91.6%, 45.5%, and 65.7% respectively. The average MLR declined from 0.212 (95% CI: 0.190 – 0.235) at baseline to 0.182 (95% CI: 0.166 – 0.198) after three months of TPT. A viral load of >50 copies/ml (aOR, 5.67 [1.12-28.60]) was associated with a high MLR while that of <50 copies/ml (aOR, 0.07 [0.007-0.832]) was associated with a decline in MLR. MLR was highly specific in diagnosing latent TB and declined significantly following three months of TPT. Implications of a high MLR and decline in MLR after TPT need further evaluation in a larger cohort.

## Background

Identification and preventive treatment of individuals with *Mycobacterium tuberculosis* (*M. tuberculosis)* infection are critical in achieving the World Health Organization’s (WHO) ambition to end the tuberculosis pandemic by 2035 [1]. The Interferon Gamma Release Assay (IGRA) has a high specificity and negative predictive value (NPV) for diagnosis of latent tuberculosis (TB), however, it’s use is limited by the requirement for sophisticated laboratory equipment, skilled human resources, and the inherent high cost of supplies required to carry out the test [2]. While on the other hand, the Tuberculin Skin Test’s (TST) cross-reactivity with the Bacillus Calmette-Guérin (BCG) vaccination and non-tuberculous mycobacteria, leads to a low specificity of TST, which has been reported as low as 57% [3]. The above factors have limited the deployment of IGRA and TST in resource-limited settings, which has led to the majority of clinicians providing TB preventive therapy (TPT) to all individuals living with HIV with no evidence of active TB, excluded by a negative WHO TB symptom screen. Moreover, it has been demonstrated that TPT is more beneficial for people who have a positive latent TB test [4], hence targeting TPT to individuals who are more likely to benefit from it would prevent unwanted exposure to TPT and be more economical.

The WHO four symptom screen with absence of cough, weight loss, night sweats, and fever excludes active TB in individuals targeted for TPT [5]. Although a meta-analysis by Hamada *et al* showed that the symptom screen had a fairly good sensitivity when used to exclude active TB in antiretroviral therapy (ART) naïve people living with HIV (89.4%), the specificity was very low (28.1%) [6]. The sensitivity was however lower (51%) in patients on ART and in other conditions like pregnancy [6]. The sensitivity in patients on ART improved to 84.6% with the addition of the chest x-ray (CXR) but at the expense of specificity, which dropped from 55.5% to 29.8% [6]. The low specificity is of particular concern because of the dire consequences of exposure to inappropriate treatment, including driving the emergence of drug resistance [6]. The limitations of the IGRA, TST and the symptom-based screen necessitate alternative tools to improve the screening for latent TB and determining response to TPT.

There is currently no bacteriological, molecular, or radiological tool used to monitor response to TPT. Clinical monitoring for TB reactivation is the only available tool because IGRA and TST may not revert to negative following TPT [7, 8], due to the persistence of effector memory cells in absence of any continuing antigen stimulation. Based on the above dilemmas, there have been some attempts to develop alternative biomarkers for the diagnosis of latent TB and monitoring response to preventive therapy. *Mycobacterium tuberculosis* DNA in peripheral blood mononuclear cells has been evaluated as a biomarker for TB infection and monitoring response to TPT. A recent study described that the proportion of participants in whom *M. tuberculosis* DNA was detectable at baseline declined by 42% following TPT [9]. Despite their promise, *M. tuberculosis* DNA-based biomarkers still need further evaluation, and like IGRA, may not easily be deployed in limited resource settings.

Monocytes replenish macrophages [10], which are the primary effector cells against *M. tuberculosis* infection [11]. In addition, through the interferon-gamma (IFN-γ) signaling, *M. tuberculosis* biases hematopoiesis towards the proliferation of the myeloid hematopoietic stem cells (HSCs) rather than lymphoid cells, which include monocytes to propagate infection [12-14]. IFN-γ signaling activates the expression of interferon regulatory factors (IRF), which promote differentiation towards the myeloid lineage [12]. This leads to a higher Monocyte to Lymphocyte Ratio (MLR) in active TB compared to latent TB or healthy donors [15, 16]. The MLR, is a rapid and affordable biomarker that has the ability to discern latent TB from active TB, given that a higher MLR is seen in adults and children with active TB compared to latent TB ([16, 17]. An MLR cut-off of 0.38 in Kenyan children living with HIV had a sensitivity of 77%, specificity of 78%, positive predictive value (PPV) of 24%, and NPV of 97%, for the diagnosis of active TB [16]. A lower cut-off (0.29) in Italian HIV-uninfected adults had high sensitivity, 91%, and specificity, 94% [16]. Besides being useful in diagnosis, the MLR was also useful in monitoring treatment of active TB in children living with HIV with confirmed TB, in whom it declined significantly from 0.41 before treatment to 0.11 after treatment [18].

We, therefore, sought to compare the performance of the MLR versus IGRA in the diagnosis of latent TB, and to measure the change in MLR after three months of TPT among Ugandan adults living with HIV, in whom TB had been excluded using a negative WHO symptom screen [5]. The results from the study could bridge the gaps in diagnosing latent TB and monitoring the response to TB preventive therapy, where resources are limited.

### Methodology

This was a prospective observational study carried out between May and November 2021 at the Infectious Diseases Institute (IDI), Makerere University in Kampala, Uganda. The current standard of care is the provision of TPT to all people living with HIV and a negative TB symptom screen. We consecutively enrolled people living with HIV of 18 years and above, with no prior TB treatment, and a negative TB symptom screen, which included exclusion of patients with a cough, weight loss, fevers, and night sweats. A thorough physical examination for respiratory system abnormalities, lymphadenopathy, enlarged liver, spleen, and other relevant clinical features to exclude those with features suggestive of TB, was also performed. Participants with a negative symptom screen and no clinical findings suggestive of TB, were consented and administered the study questionnaire. The sample size was derived using the formula for diagnostic studies and a sample size of 105 participants gave an 80% chance of delineating the diagnostic performance of the MLR versus IGRA in diagnosing latent tuberculosis. The final sample size was 115, considering a 10% loss to follow up.

Individuals living with HIV were invited to give written consent to participate in the study after being briefed about the study. Consenting participants gave social demographic information, which was collected using the study questionnaire. Only authors who were part of the recruiting study team had access to the patient personal identifying data, which was kept as logs separate from the case record forms, under double local storage system. Information was collected on history of contact with an index pulmonary TB patient, smoking status, alcohol intake, BCG immunization status, and last HIV viral load results. Formal employment, private business, casual laborer, peasant, farmer, and others were the subcategories assessed for occupation. Given the need to have higher-powered subcategory numbers, the above were re-categorized according to the risk for *M. tuberculosis* infection, including congestion and other risky working and living conditions like ventilation and exposure to silica. Due to comparable risks posed, formal employment and private business were categorized as non-casual employment, while the rest were classified as casual employment or unemployment. Education was categorized as no education, primary and post primary education. To minimize bias, recruitment was spread over three months, calibrated instruments used to measure weight and height, and only the question on history of contact with an index pulmonary TB patient required remote recall. The outcome variables included a high or low MLR and a positive or negative IGRA.

Isoniazid 300 mg plus 25 mg pyridoxine daily for six months was initiated after intensive adherence counselling. Participants received three monthly drug refills and adherence support was provided through daily reminders by the next of kin and monthly phone call reminders by the study team.

Participants donated 5 ml of blood, of which 4 ml was used for IGRA and 1 ml for baseline complete blood count (CBC) testing. Another 1 ml of blood was collected after three months of TPT for the follow-up CBC. Absolute monocyte and lymphocyte counts were derived from these to calculate the MLR. The CBC was analyzed immediately using the Beckman Coulter Ac•T 5diff AL (Autoloader) Hematology Analyzer. Blood for IGRA testing was collected at baseline in specialized QuantiFERON tubes and transported to the laboratory at room temperature within 16 hours. The IGRA samples were incubated at 37°C for 16 to 24 hours, centrifuged at 3000g for 10 minutes, and the serum stored at -80°C until analysis, which followed the standard operating procedure provided by the Qiagen Company, the makers of the QuantiFERON kits. A positive IGRA was taken as a TB1 or TB2 antigen value minus the Nil sample value ≥ 0.35 and ≥ 25% of Nil and vice versa for the negative IGRA if the Mitogen value was ≥ 0.5, and if it was < 0.5 the result was taken to be intermediate. The IGRA and CBC tests were performed in the IDI Translation Research Laboratory and the IDI Makerere University-John Hopkins University (MUJHU) laboratory respectively.

Ethical approval was obtained from the Makerere University School of Biomedical Sciences Institutional Review Board, reference number SBS 794. Written informed consent was obtained from all participants before any study-related procedures were conducted. The study did not include minors or vulnerable groups.

### Statistical analysis

For descriptive analysis, continuous variables were reported as means with standard deviation (SD) for normally distributed data or median and interquartile range (IQR) for non-normally distributed data while categorical variables were reported as proportions in terms of frequencies and percentages. The MLR was derived from the ratio of the absolute monocyte count to the absolute lymphocyte count while the diagnostic cut-off for MLR was determined using the discriminant function analysis performed in excel. A function was generated from a sample of known positive and known negative cases based on the IGRA test. The function was then applied to the results of test diagnostic test (MLR) to classify them as positive or negative. Results of the discriminant analysis displayed a discriminant score and predicted group membership for each case. A value lower than the cut-off score represented a low MLR while the one above represented a high MLR. IGRA was used as the gold standard for diagnosis of latent TB. The sensitivity, specificity, PPV and NPV were determined from a 2X2 table of MLR and IGRA. A receiver operator curve (ROC) curve was plotted using sensitivity (y-axis) and 1-sensitivity (x-axis). A paired sample t-test was used to determine whether there was a statistically significant difference between mean MLR at baseline and that at follow up, while, a multivariate logistic regression analysis with forward stepwise logit function was employed to identify independent correlates of a high MLR and decline in MLR. Age, gender, BCG scar, viral load, body mass index (BMI), and IGRA status were included in the regression analysis based on the biological plausibility. A P<0.05 was considered statistically significant. Missing data including that, which was due to loss to follow up, was not included in the analysis.

## Results

### Study participant characteristics

Participant socio-demographic characteristics are shown in Table 1. Of the 110 people living with HIV recruited, 71.8% (79/110) were female and had a median age of 43 years (interquartile range (IQR), 30 - 51). The majority of the participants (49/110, 44.6%) were over the age of 45 years. Of these participants, 73.6% (81/110) had a BCG scar - suggesting previous TB vaccination and 82.5% (85/110) had a suppressed viral load. All participants were receiving ART and they all received isoniazid for six months but the follow-up CBC measurement was at three months. Twenty-one percent (23/110) of participants did not complete follow-up and therefore did not have follow-up CBC data.

**Table 1:** Socio-demographic characteristics.

| Variable | Frequency | Percentage (%) |
| --- | --- | --- |
| Age (years) |  |  |
| **Below 25** | 16 | 14.55 |
| **25-45** | 45 | 40.91 |
| **>45** | 49 | 44.55 |
| **Education** |  |  |
| **No education** | 4 | 3.67 |
| **Post Primary** | 70 | 64.22 |
| **Primary** | 35 | 32.11 |
| **Employment** |  |  |
| **Non casual** | 57 | 51.82 |
| **Casual** | 53 | 48.18 |
| **Alcohol** |  |  |
| **No** | 87 | 79.09 |
| **Yes** | 23 | 20.91 |
| **Smoking history** |  |  |
| **Never smoked** | 106 | 96.36 |
| **Current smoker** | 4 | 3.64 |
| **BCG Status** |  |  |
| **No** | 29 | 26.36 |
| **Yes** | 81 | 73.64 |
| **IGRA test** |  |  |
| **Negative** | 71 | 64.55 |
| **Positive** | 39 | 35.45 |
| **HIV Viral load (copies/ml)** |  |  |
| **<50** | 85 | 82.52 |
| **>50** | 18 | 17.48 |

### Diagnostic performance

We derived an MLR cut-off of 0.35 for the diagnosis of latent TB, indicating the presence of latent TB infection in individuals who scored above this value, i.e. high MLR. Based on this cut-off, the sensitivity, specificity, PPV, and NPV were 12.8% (95% CI: 6.6% - 19.1%), 91.6% (95% CI: 86.4% - 96.8%), 45.5% (95% CI: 36.2% - 54.8%), and 65.7% (95% CI: 56.8% - 74.5%) respectively. The area under the curve was 0.45, *figure 1*.

**Figure 1:**
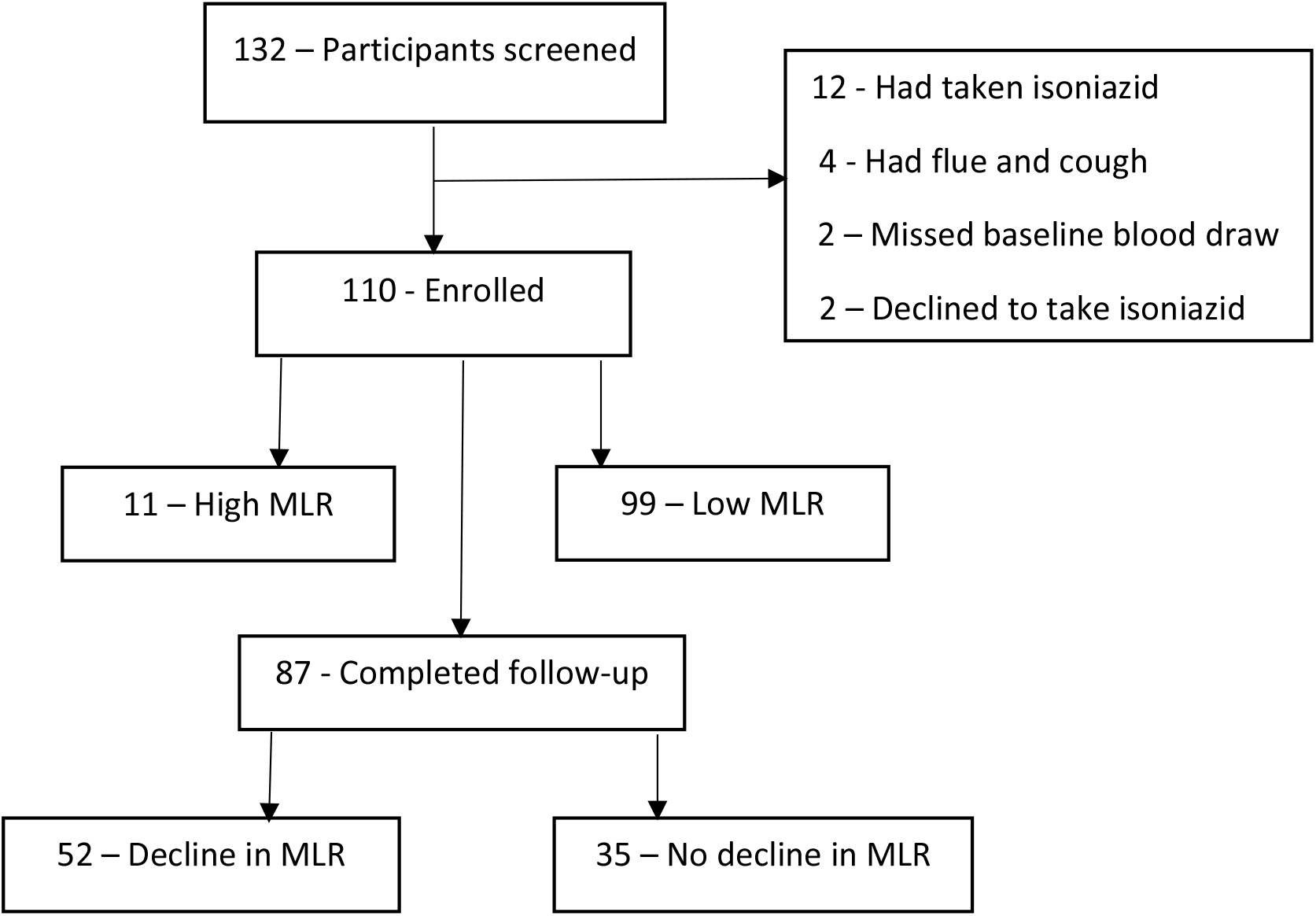
Study flow chart

**Figure 1:**
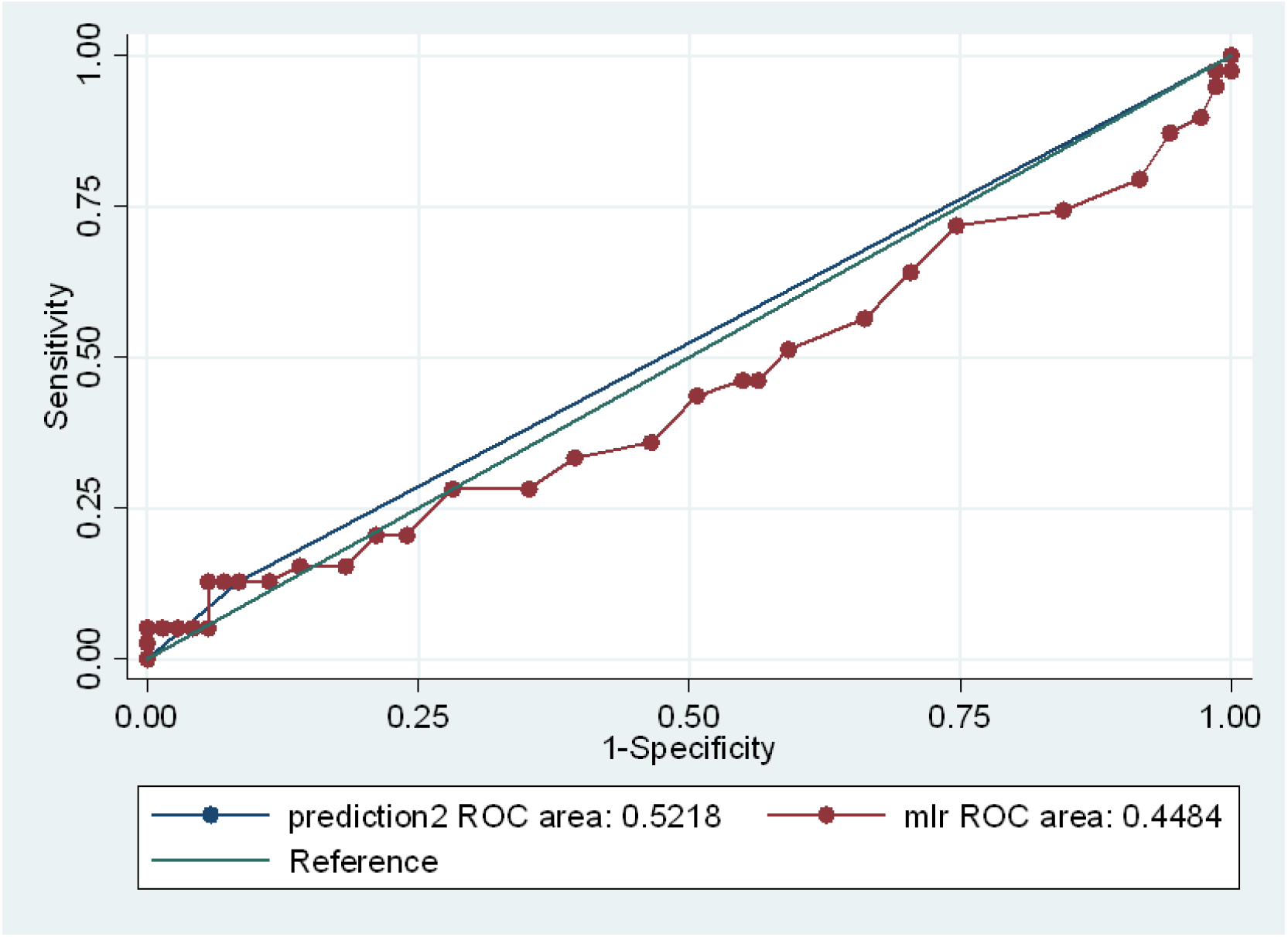
The Receiver Operative Curve for the diagnostic utility of MLR.

### Effect of 3-month IPT on MLR

A total of 87/110 participants with both baseline and three month follow-up MLR results were included in the analysis of the effect of isoniazid-based TPT on MLR. The average MLR for all participants significantly decreased from 0.212 (95% CI: 0.190 – 0.235) at baseline to 0.182 (95% CI: 0.166 – 0.198) at 3 months of TPT, representing a decline of 0.030 (95% CI: 0.007 – 0.053), *table 2*. We found that 52/87 (59.8%) of the participants had a decrease in MLR following three months of TPT, majority of which were >45 years (27/52, 51.9%), were female (35/52, 67.3%), had a BCG scar (39/52, 75.0%), were IGRA negative (34/53, 65.4%), and had a viral load of < 50 copies/ml (41/48, 85.4%), *table 4*.

**Table 2:** Effect of IPT on MLR.

| Variable | Obs | Mean | Std. Err. | Std. Dev. | [95% Conf. Interval] |
| --- | --- | --- | --- | --- | --- |
| Baseline MLR | 87 | 0.2121839 | 0.011318 | 0.105571 | 0.1896837 0.234684 |
| Follow-up MLR | 87 | 0.182069 | 0.007846 | 0.073186 | 0.166471 0.197667 |
| Difference | 87 | 0.0301149 | 0.011742 | 0.109518 | 0.0067735 0.053456 |

**Table 3:**
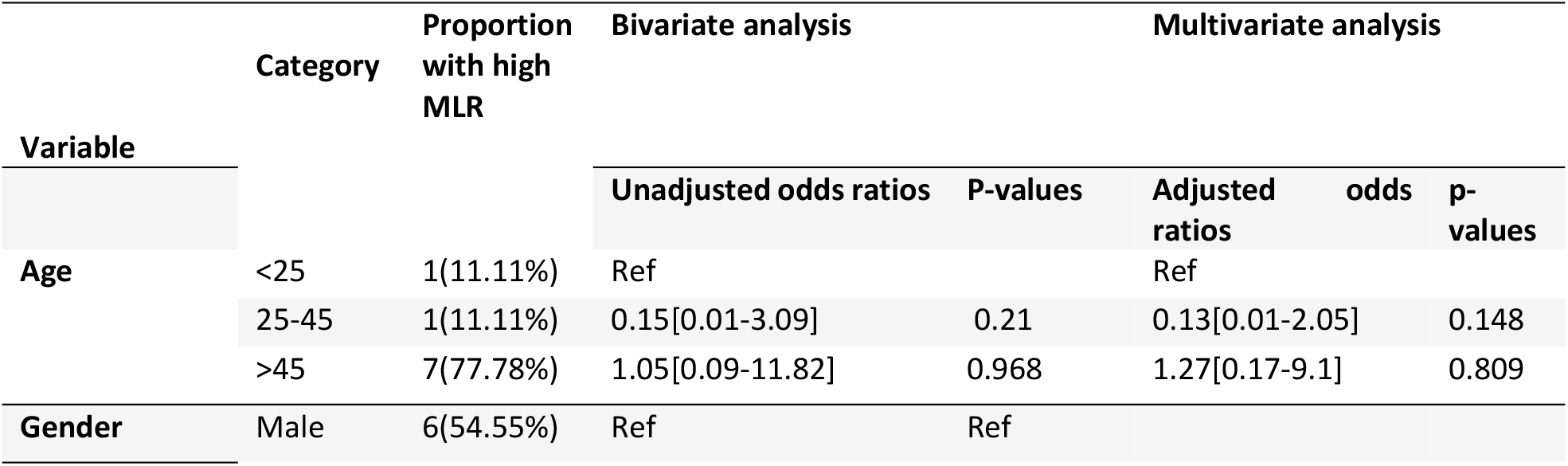
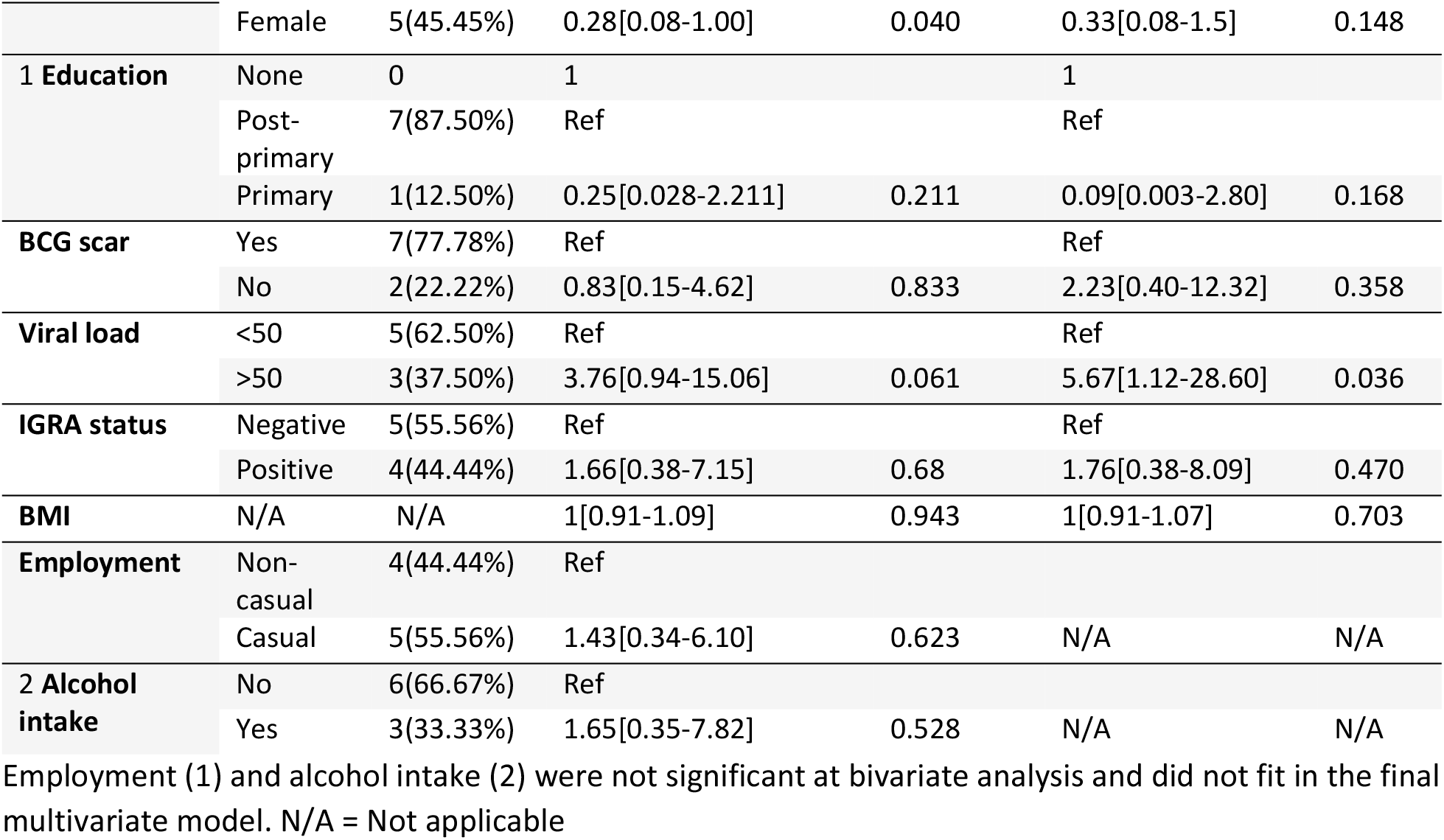
Factors associated with a high MLR.

**Table 4:** Factors associated with a decline in MLR

| Variable | Category | Proportion with decline in MLR | Bivariate analysis |  | Multivariate analysis |  |
| --- | --- | --- | --- | --- | --- | --- |
|  |  |  | Unadjusted odds ratios | P-values | Adjusted odds ratios | p-values |
| **Age** | <25 | 21 (40.38%) | Ref |  | Ref |  |
|  | 25-45 | 4 (7.69%) | 0.15[0.01-3.09] | 0.21 | 0.2[0.015-2.65] | 0.222 |
|  | >45 | 27 (51.92%) | 1.05[0.09-11.82] | 0.968 | 1 |  |
| **Gender** | Male | 17 (32.69%) | Ref |  | Ref |  |
|  | Female | 35 (67.31%) | 0.31[0.071-1.352] | 0.119 | 0.28[0.04-2.04] | 0.210 |
| **BCG scar** | Yes | 39 (75.00%) | Ref |  | Ref |  |
|  | No | 13 (25.00%) | 0.83[0.15-4.62] | 0.833 | 1.28[0.13-12.93] | 0.836 |
| **Viral load** | <50 | 41 (85.42%) | 0.19[0.03-1.08] | 0.061 | 0.07[0.007-0.832] | 0.035 |
|  | >50 | 7 (14.58%) | Ref |  |  |  |
| **IGRA status** | Negative | 34 (65.38%) | Ref |  | Ref |  |
|  | Positive | 18 (34.62%) | 1.66[0.38-7.15] | 0.68 | 0.79[0.09-6.59] | 0.828 |
| **BMI** | N/A | N/A | 1[0.91-1.09] | 0.943 | 1.04[0.93-1.16] | 0.524 |
| **1 Education** | None | 2 (3.92%) | 1 |  | N/A | N/A |
|  | Post-primary | 33 (64.71%) | Ref |  | N/A | N/A |
|  | Primary | 16 (31.37%) | 0.25[0.028-2.211] | 0.211 | N/A | N/A |
| 2<br>**Employment** | Non-casual | 25 (48.08%) | Ref |  | Ref |  |
|  | Casual | 27 (51.92%) | 1.43[0.34-6.10] | 0.623 | N/A | N/A |
| 3 **Alcohol intake** | No | 39 (75%) | Ref |  | N/A | N/A |
|  | Yes | 13 (25%) | 1.65[0.35-7.82] | 0.528 | N/A | N/A |

Education (1), employment (2), and alcohol intake (3) were not significant at bivariate analysis and did not fit in the final multivariate model. N/A = Not applicable

### Factors associated with a high MLR at baseline and declining MLR

At bivariate analysis, only female gender (unadjusted odds ratio (OR), 0.28 [0.08-1.00]) was associated a high MLR, while, only an HIV viral load of >50 copies/ml (adjusted OR (aOR, 5.67 [1.12-28.60]) was associated with a high MLR at multivariate analysis, *table 3*. Only an HIV viral load of <50 copies/ml (aOR, 0.07 [0.007-0.832]) was associated with a decline in MLR, at multivariate analysis, *table 4*.

## Discussion

We have demonstrated a high specificity (91.6%) of MLR for diagnosis of latent TB using a cut off of 0.35, which is comparable to IGRA’s (90%) [19] in people living with HIV. Based on this cut off, MLR had a lower sensitivity than IGRA (12.8% vs 64%) [20] in people living with HIV. To our knowledge, this is the first study demonstrating a low sensitivity but high specificity of MLR in diagnosing latent TB. Although MLR’s limited sensitivity would prevent it from being used as a major diagnostic test for latent TB, its high specificity positions it as an inexpensive and simple biomarker that could be used to improve TB screening with the WHO symptom screen.

The ability of MLR to monitor response to TB treatment has been observed in active TB, where there was a significant decline in MLR after six months of TB therapy and a persisting high MLR was associated with treatment failure [21]. We have demonstrated for the first time that MLR declined during TPT in the majority of participants. This could lead to a paradigm shift in the monitoring of response to TPT. Currently, there is no laboratory or radiological test to monitor response to TPT. Given that TST and IGRA have low reversion rates following TPT [22, 23], clinical absence of active TB during and after TPT is currently the measure of effectiveness of TPT. Our results suggest that MLR is a potential TPT monitoring biomarker that can overcome the limitations of the current tests.

Among the factors studied, only HIV viral load was associated with a high MLR and a decline in MLR after TPT. Participants with viral loads >50 copies/ml had a higher risk of having a high baseline MLR than those with a viral load of <50 copies/ml, while, participants with a viral load of <50 copies/ml were more likely to have a decline in MLR than those with a viral load >50 copies/ml. Due to the impaired immunological response towards opportunistic infections, ongoing HIV replication, as indicated by a high viral load, is an independent risk factor for *M. tuberculosis* infection [24]. On the other hand, a suppressed viral load is linked to immunological restoration and improved bacilli burden control, which aids response to TPT [25].

Our study had some limitations. First, the sample size was powered to study the diagnostic utility of the MLR and not the factors associated with a high MLR. This could have limited our ability to clearly describe factors associated with a high baseline MLR. Secondly, only 87 of the 110 participants had a complete pair of baseline and follow-up MLR, which may have affected the evaluation of the factors associated with a decline in MLR. Thirdly, the effect of IPT was assessed after 3-months of isoniazid-based TPT rather than six months, which may have underestimated the effect of TPT on the MLR. It also still needs to be clarified what impact the one-month TPT regimen would have on the MLR response.

## Conclusion

The MLR had high a specificity in the diagnosis of latent TB and declined significantly in the majority of patients following TPT. These data if validated in larger cohorts, could improve the screening of latent TB using the WHO four symptom screen and avail a biomarker that could monitor the response to TPT.

## Data Availability

Data will be availed after acceptance

## Recommendation

Larger studies are needed to further evaluate the effect of TPT on MLR and the implication of a high MLR before and after TPT.

## Author contribution

Jonathan Mayito, Conceptualization, Investigation, Methodology, Project Administration, Writing – Original Draft Preparation

David B. Meya, Conceptualization, Investigation, Methodology, Resources, Supervision, Writing – Review & Editing

Miriam Akia, conduct of the study procedures, Writing – Review & Editing

Flavia Dhikusooka, statistical analysis, Review and Editing

Joshua Rhein, Conceptualization, Investigation, Methodology, Supervision, Writing – Review & Editing

Christine Sekaggya-Wiltshire, Conceptualization, Investigation, Methodology, Supervision, Writing – Review & Editing

